# Using kinematics to re-define the pull test as a quantitative biomarker of the postural response

**DOI:** 10.1101/2021.05.24.21257163

**Authors:** Samuel Daly, Jacob T. Hanson, Vibha Mavanji, Amy Gravely, Scott Lewis, James Ashe, John M. Looft, Robert A. McGovern

## Abstract

**Background:** Quantitative biomarkers are needed for the diagnosis, monitoring and therapeutic assessment of postural instability.

**Objective:** Create a practical and objective measure of postural instability using kinematic measurement of the pull test to incorporate into clinical practice.

**Methods:** Twenty patients evaluated and treated for normal pressure hydrocephalus were tested over a number of sessions. Patients were fitted with 15 inertial measurement units during each session. At each session, the patient underwent 10-20 pull tests performed by a trained clinician. The clinician purposefully induced a range of perturbations during each session to assess the patient’s step response. Kinematic data was extracted for each pull test and aggregated.

**Results:** Patients participated in 57 sessions for a total of 860 trials and were separated into groups by pull test score. The center of mass velocity profile easily distinguished between groups such that score increases correlated with decreases in peak velocity and later peak velocity onset. All patients except those scored as “3” demonstrated an increase in step length and decrease in reaction time with increasing pull intensity. Groups were distinguished by differences in overall step length or reaction time regardless of pull intensity (y-intercept). A logistic regression model including only kinematic variables identified three kinematic factors that could be used to identify trials in which patients needed to be caught (i.e. “fell”).

**Conclusion:** An instrumented, purposefully varied pull test produces kinematic metrics useful for distinguishing clinically meaningful differences between and within NPH patients. These metrics should be followed prospectively to examine fall risk.

## Introduction

Balance impairments are common in older adults and lead to falls with subsequent injuries, reduced mobility, and decreased quality of life^1^. In neurodegenerative disorders such as Parkinson’s Disease (PD) and normal pressure hydrocephalus (NPH), balance impairment – or postural instability – is a key feature of the disease process and can be defined as an inability to maintain an upright posture in response to perturbations or changes in the environment. Neurologists and neurosurgeons commonly assess postural instability using the pull test (UPDRS_PT_)^2^ or push and release test^3^, while physiatrists and physical therapists may also use the Berg Balance Scale,^4^ Mini-BEST^5^ or a combination of all of these assessments. While useful to individual providers, these assessments are based on semi-objective ordinal ratings and are therefore subject to variability in both execution and interpretation^6,7^. The variety of testing and subjective scoring can make effective communication between providers and specialties difficult when discussing a particular patient’s condition.

Quantitative biomarkers for postural instability have historically required a large, dedicated setup and are time-consuming, costly, and not widely available to clinicians. Thus, identifying easily implemented biomarkers for the diagnosis, monitoring and therapeutic assessment of postural instability in the clinical setting is a priority.^8^ Such a biomarker should be able to accurately communicate a patient’s condition between providers and standardize clinical trial outcomes by negating any dependence on interrater reliability. Improvements in wearable technology have led to the emergence of these devices as potential tools in the development of quantitative biomarkers for gait and postural instability^9,10^. Metrics from wearable sensors have been developed for almost all aspects of balance including quiet standing^11,12^, anticipatory postural adjustments^13^, and balance during gait^14–16^. The major challenges with wearable technology are relating these developed metrics to clinically meaningful results such as fall risk and identifying which metrics are specific to different disease processes. For example, patients with PD will likely have a different pattern of deficits on quantitative wearable metrics compared to patients with NPH or cerebellar ataxia. This will depend on both the underlying disease process and the metrics chosen to evaluate patients.

In this study, we present a method of using kinematic measurements from the pull test in NPH patients to create a quantitative, reliable, and clinically useful measure of postural instability. We have chosen to quantify the UPDRS_PT_ because it is the neurological gold standard and most widely used clinical test available for postural instability. Kinematics examines the features of the body in motion without regard for the forces which caused the motion. Although multiple complex motor control systems may be involved in the production of a postural response, the UPDRS_PT_ allows the clinician to assess the ultimate response. Thus, as an approach focused on outcomes, kinematics lends itself to the quantitative clinical translation of the UPDRS_PT_.

## Methods

### Subjects

Twenty patients were consecutively prospectively enrolled over a period of 18 months from the Minneapolis VA Health Care System (MVAHCS). All patients were diagnosed with either possible or probable NPH and referred for neurosurgical evaluation. They were excluded if they did not have the capacity to consent as identified by the University of San Diego Brief Assessment of Capacity to Consent (UBACC). Patients who were identified to have other forms of parkinsonism were also excluded. We collected relevant demographic data from each patient as well as their responses to a number of questionnaires related to their balance and falls. Patients underwent kinematic, neuropsychological and physical therapy assessment pre- and post-lumbar drain placement. The treating neurosurgeon (R.M.) used these assessments to decide whether to offer treatment with ventriculoperitoneal shunt (VPS) placement. Regardless of the surgical decision, all patients were prospectively followed with continued assessments. This study was approved by the MVAHCS Institutional Review Board, and all patients provided informed consent for participation.

### Task Details

Participants were equipped with a set of 15 inertial measurement units (IMUs) during each pull test session (Xsens, En Schede, Netherlands). The pull test was executed for each patient by a trained clinical examiner (either a practicing movement disorders neurologist or neurosurgeon). The examiner followed the instructions on the UPDRS form for conducting the pull test^2^ and conducted between ten and twenty pull test trials for each patient. The examiner used clinical discretion to determine the force of the induced perturbation during the recorded trials, but they were instructed to use a variety of intensities throughout the trials as able. The first trial after the instructional trial was scored in the standard manner on the MDS-UPDRS scale.

### Data and Statistical Analysis

Center of mass (COM) and foot position data were exported from the recorded motion capture analysis file and imported and analyzed within Igor Pro 6.00 (Wavemetrics, Oregon, USA) to calculate velocity and acceleration for the COM, feet and other body segments. Custom functions were then used to identify the relevant points of interest (Supplemental Figure 1). Once relevant points were extracted from the trials, they were exported into R (v 3.6.2) ^18^and further analyzed. Plots were created using the R package “ggplot2.” Supplemental Table 1 contains a description and interpretation of the relevant kinematic variables while Supplemental Figure 1 demonstrates the relevant points of interest for each individual trial^17^.

Statistically significant differences in center of mass velocity (V_COM_) plots were considered to be non-overlapping 95% confidence intervals of timeframes 50 ms or greater. Linear models were created to analyze the interaction between step length and pull intensity as well as reaction time and pull intensity for each UPDRS_PT_ group. To test for an overall effect, we first performed ANCOVA. If the effect was significant, we then compared differences between the slope and intercept of each of these groups using pair-wise comparisons with a Tukey HSD correction. For fall trial analysis, we compared mean values of relevant kinematic variables using two-sided student’s t-tests. A binary logistic regression model was created to assess the effects of kinematic variables on the likelihood of a “fall” occurring. Interaction terms were included for variables known to have significant association with one another (e.g., step length and pull intensity). Odds ratios for each variable and their interactions were calculated along with 95% confidence intervals. P values less than 0.05 were considered statistically significant.

## Results

### Participants

We tested 20 patients diagnosed with possible or probable NPH over a period of 18 months. They participated in 57 sessions with a mean of 15.1 pull test trials per session, for a total of 860 trials. Demographic data can be found in Table 1. Patients were grouped by UPDRS_PT_ scores for stratified analysis.

**Table 1.** Patient Demographics

| Patient ID | Age Range | Sex | Height (cm) | Weight (kg) | Treated with VP Shunt? | Number of Pull Test Sessions | Number of Pull Test Trials | Mean UPDRS<sub>PT</sub> Score | Mean ABC Score |
| --- | --- | --- | --- | --- | --- | --- | --- | --- | --- |
| 1 | 65-69 | M | 183 | 111.4 | N | 2 | 24 | 1.70 | 81.3 |
| 2 | 75-79 | M | 173 | 98.6 | Y | 6 | 93 | 0.38 | 93.8 |
| 3 | 75-79 | M | 159.5 | 78.6 | Y | 6 | 97 | 1.22 | 76.3 |
| 5 | 60-64 | M | 171 | 100 | Y | 5 | 81 | 0.83 | 78.1 |
| 6 | 75-79 | M | 161 | 89.5 | Y | 5 | 88 | 1.14 | 78.1 |
| 7 | 80-84 | M | 153.2 | 59.4 | Y | 4 | 34 | 2.83 | 9.4 |
| 8 | 70-74 | M | 175 | 98.2 | N | 2 | 28 | 2 | - |
| 9 | 80-84 | M | 184.2 | 114.1 | Y | 4 | 60 | 1.83 | 79.1 |
| 10 | 80-84 | M | 182 | 79.3 | Y | 4 | 66 | 2.67 | 56.3 |
| 12 | 70-74 | M | 177 | 79.2 | Y | 3 | 53 | 1.60 | 78.8 |
| 13 | 55-59 | M | 185 | 141 | Y | 4 | 63 | 0.90 | 89.1 |
| 15 | 65-69 | M | 163 | 70.5 | N | 2 | 27 | 3 | 10.3 |
| 16 | 75-79 | M | 162 | 69.5 | N | 2 | 29 | 2.67 | 95.9 |
| 17 | 65-69 | F | 178 | 82.7 | Y | 2 | 32 | 1.83 | 64.1 |
| 18 | 75-79 | M | 175 | 64.1 | N | 2 | 30 | 1 | 86.3 |
| 19 | 65-69 | M | 184 | 102.4 | Y | 2 | 31 | 0.83 | 47.2 |
| 20 | 75-79 | M | 171 | 76.3 | N | 2 | 24 | 2.5 | 93.1 |
| Mean (sd) | 72.8 (7) | 19M:1F | 172.8 (9.9) | 89.1 (21) | 14Y:6N | 3.35 (1.5) | 50.6 (26) | 1.70 (0.8) | 69.8 (27) |

Abbreviations: UPDRS<sub>PT</sub> = Unified Parkinson's disease Rating Scale for Pull Test. VP = Ventriculoperitoneal. ABC = Activities-Specific Balance
Confidence Scale.

### V_COM_ Profile Differentiates between UPDRS_PT_ Scores

We first sought to describe an objective, kinematic profile for each pull test trial using the patient’s center of mass velocity (V_COM_). In its simplest terms, each pull test trial can be described by the patient’s V_COM_ as it begins at zero when the patient is at rest, rises as the patient is pulled backwards, continues to rise as the patient steps backwards and then declines and approaches zero as the patient recovers from the perturbation (Supplemental Figure 1). When we stratified each patient session by UPDRS_PT_ score and averaged V_COM_ for all trials, we found statistically significant differences in V_COM_ between all groups of patients within the first 500 ms after pull onset. Increases in UPDRS_PT_ score correlated with decreases in peak V_COM_ values and later peak V_COM_ onset indicating that V_COM_ profiles discriminated between clinically meaningful criteria (Figure 1A).

**Figure 1.**
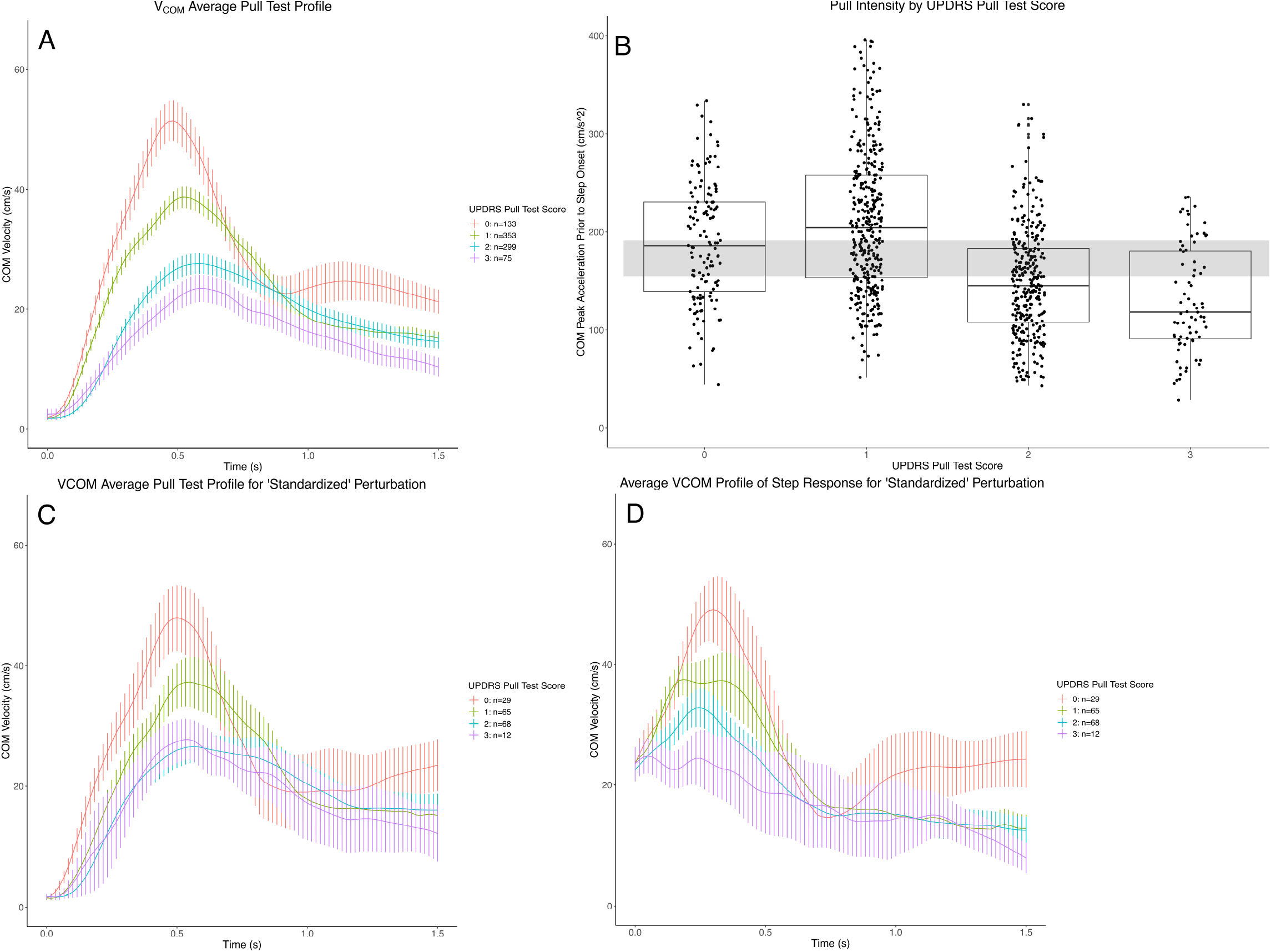
A) Aggregate V_COM_ profiles over time stratified by UPDRS_PT_ score. B) Representation of identifying “standardized” pull tests. C and D) Aggregate V_COM_ profiles over time for a “standardized” perturbation stratified by UPDRS_PT_ following the: C) perturbation, and D) step onset. Abbreviations: UPDRS_PT_ = Unified Parkinson’s Disease Rating Scale for the Pull Test. V_COM_ = Center of Mass Velocity (VCOM).

### Examining the Step Response to a “Standardized” Perturbation

We considered that some of the difference in V_COM_ profiles were a result of intentionally inducing perturbations of varying intensities, as patients who were pulled harder likely would demonstrate higher peak V_COM_ during their trials. Patients scored as “0”(188.5 ± 61 cm/s) or “1” (209.5 ± 72 cm/s) were pulled harder on average than patients scored as “2” (148.7 ± 56 cm/s) or “3”(130.5 ± 54 cm/s) (Figure 1B, Supplemental Table 2). Therefore, we next restricted our analysis to a narrow range of pull intensities (40^th^ to 60^th^ percentile, shaded area in Figure 1B), as measured by peak A_COM_ prior to step onset (see Supplemental Methods and Supplemental Table 1 for details). This changed the V_COM_ profiles in a few ways (Figure 1C). First, patients scored as “0” had lower peak V_COM_ values, reducing the differences seen between “0” and all other groups. Second, patients scored as “3” had a slightly higher mean peak V_COM_ and a slightly reduced time to peak V_COM,_ eliminating much of the differences in V_COM_ profiles between groups “2” and “3.” All other groups’ peak V_COM_ values and time to peak V_COM_ stayed essentially the same. Statistically significant decreases in peak V_COM_ as well as later peak V_COM_ onset remained associated with increasing UPDRS_PT_ scores except between the patients scored as “2” and “3”. Statistically significant differences in V_COM_ remained between groups “0”, “1” and the other two groups during the 200-500 ms time period (Figure 1C). This time period coincides with the initiation of step onset for patients.

Since groups “0” and “1” were still able to be differentiated from groups “2” and “3” within this narrow range of perturbations (particularly during the step initiation phase), we then hypothesized that differences in the step response were responsible for differentiating between groups. To examine this, we looked at patients’ step responses over this narrow range of perturbations by restricting V_COM_ exclusively to the time from step onset until the end of the trial (Figure 1D). This demonstrated the step response began for all groups at similar V_COM_ values (indicating that the perturbations were approximately standardized), but diverged within the first 300-400 ms (approximately the duration of the first step) and then converged again at the end of the second step at 700-800 ms. Statistically significant differences between similarly scored groups (i.e. “0” vs “1”, “1” vs “2”) only occurred at or near peak V_COM_ values while profiles for dissimilar groups (i.e. “0” vs. “2”, “1” vs. “3”) remained easily differentiated.

### Step Response Scaling Differentiates between UPDRS_PT_ Scores

The previous data suggests that the step responses differed by UPDRS_PT_ score, so we next investigated the step response for each individual trial in detail. We first investigated the relationship of the initial step length to the pull intensity (peak A_COM_) for each trial (Figure 2A). When separated into groups according to UPDRS_PT_ score, all groups except those classified as “3” increased their initial step length for increases in pull intensity (non-zero slope, Figure 2A, Supplemental Table 3). Groups could be stratified according to both their overall step length for a given pull intensity (y-intercept) and their ability to scale the initial step length to pull intensity (slope). In general, however, groups were best differentiated statistically according to their overall step length values for a given pull intensity (intercept). Patients scored as “0” had the highest intercept values and were statistically significantly larger than all other groups. This pattern continued for increasing UPDRS_PT_ scores with patients scored as “1” and “2” having significantly lower intercept values. Patients scored as “3” had similar intercept values to patients scored as “1” (11.2 ± 7.4 cm vs 14.5 ± 4.1 cm), but could be easily differentiated by their slope values as “3” patients were essentially unable to scale their step length to increases in pull intensity (i.e. slope = 0).

**Figure 2.**
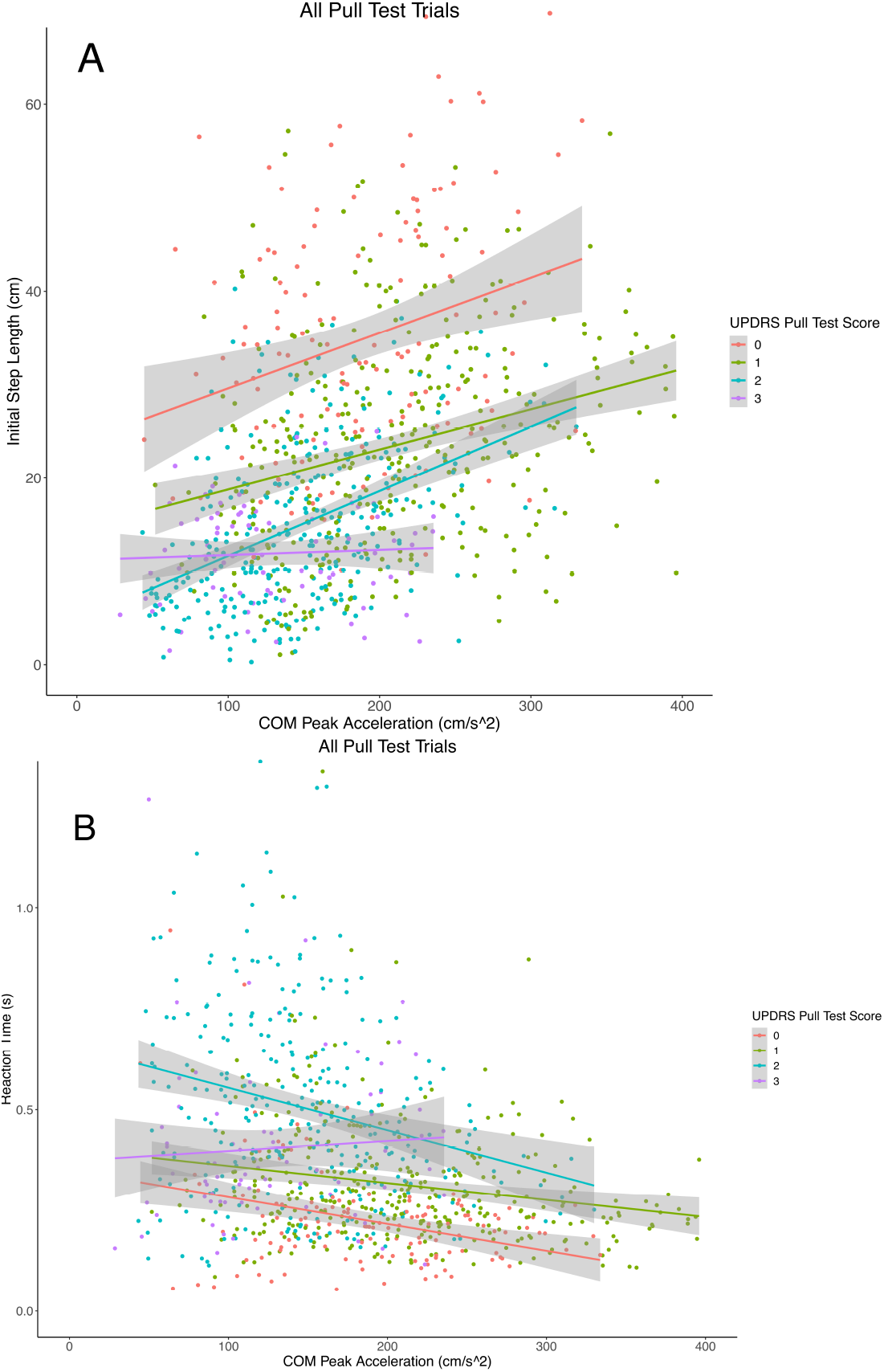
A) Initial step length as a function of the perturbation intensity, where the y-intercept represents the baseline step and the slope represents the ability to scale the step length. B) Reaction time as a function of the perturbation intensity. Abbreviations: UPDRS_PT_ = Unified Parkinson’s Disease Rating Scale Pull Test score. COM = Center of Mass.

### Reaction Time Differentiates Between UPDRS_PT_ Scores

Given the differences between groups in peak V_COM_ onset seen in Figure 1A, we hypothesized that there may be differences in the relationship of reaction time to pull intensity between them as well (Figure 2B). Similar to step length, all groups except those scored as “3” could scale their reaction time to increases in pull intensity (Supplemental Table 4). Thus, patients scored as “0”, “1”, and “2” all demonstrated quicker reaction times for increasing pull intensities (non-zero slope) while patients scored as “3” did not demonstrate any change in reaction time with increasing pull intensity (slope = 0). In general, the relationship between reaction time and pull intensity did not differentiate groups as well as the relationship between step length and pull intensity. Similar to step length, groups could mainly be distinguished by differences in the overall reaction time for a given pull intensity (y-intercept values). The fastest overall reaction times were seen in patients scored as “0” regardless of pull intensity with progressively larger intercept values (slower reaction times) with increasing UPDRS_PT_ score for patients scored as “1” or “2”. Patients scored as “3” had similar reaction time intercept values as patients scored as “0” or “1” and were defined by their inability to decrease their reaction time to increases in pull intensity (i.e. slope = 0). Groups were generally statistically indistinguishable when comparing reaction time slope with the sole exception of the comparison between patients scored as “2” and “3” (Supplemental Table 4).

### Analysis and Prediction of Falls

Patients needed to be caught by the examiner, and can therefore be considered to have “fallen,” on 8.1% of all trials. “Falls” were concentrated in four of the most severely affected patients, but they occurred in 12/20 patients overall (Supplemental Figure 2A). No patients scored as “0” fell, with an increasing proportion of falls occurring as UPDRS_PT_ score increased (Supplemental Figure 2B). Thus, we decided to investigate the kinematic relationships involved in determining whether a patient would “fall” during a pull test trial. Trials in which patients “fell” were characterized by slightly reduced and delayed peak V_COM_ values that remained close to peak V_COM_ for approximately 800 ms longer than non-fall trials (Figure 3A). This indicated patients were unable to successfully recover from the pull as they continued to move backwards without slowing down.

**Figure 3.**
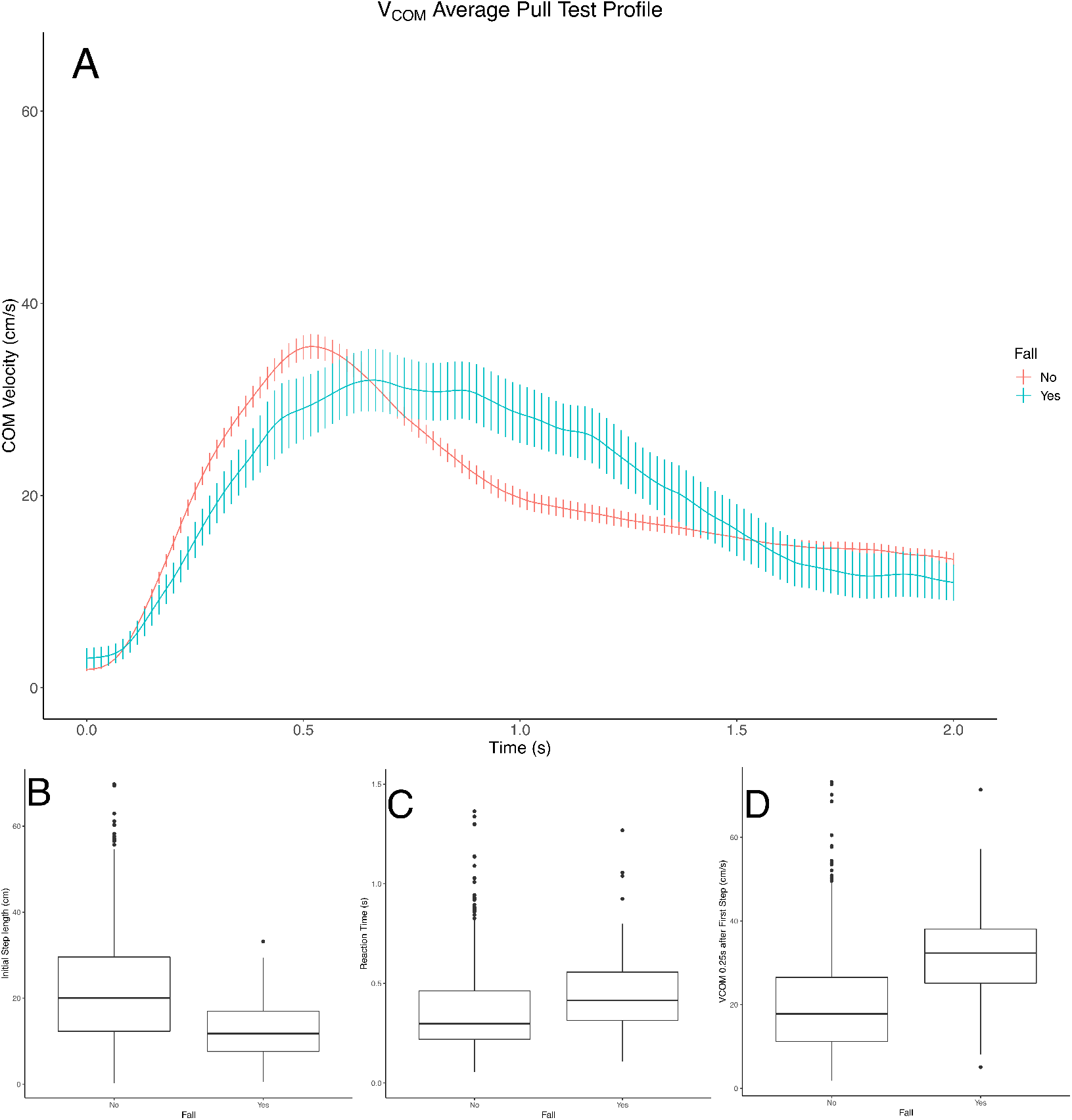
A) Aggregate V_COM_ profiles stratified by whether the patient fell. B-E) Kinematic variables associated with whether the patient fell: B) Initial step length, C) Reaction time, D) V_COM_ 0.25s after first step, and E) Time to zero velocity. Abbreviations: COM = Center of mass. V_COM_ = Center of mass velocity.

Kinematically, “fall” trials demonstrated smaller overall initial step lengths and slower reaction times (Figures 3B-C) along with other kinematic values that reflected the delayed return of V_COM_ to zero (time for V_COM_ to return to zero, V_COM_ at 0.25 s after first step offset, Figures 3D-E). In order to determine which factors were most associated with “fall” trials, we next created a logistic regression model that included only kinematic variables and their interactions. V_COM_0.25 s after first step offset, step length and pull intensity were the statistically significant factors associated with a “fall” occurring during a specific trial (whole model test, R^2^ = 0.32, p<0.0001, Table 2). To better visualize the association between V_COM_, step length and the risk of falling, we plotted the percentage of trials in which patients fell against both step length and V_COM_ at 0.25s after foot landing (Figures 4A-B). Patients who took steps greater than approximately 35 cm never fell. As step length decreased, the risk of falling increased in a non-linear fashion (Figure 4A). Similarly, if patients’ V_COM_ at 0.25s after foot landing was below 10 cm/s, their risk of falling was zero. As V_COM_ increased, their risk of falling increased (Figure 4B). The interaction of these two variables and their effect on the risk of falling is best illustrated in Figure 4C. This demonstrates that a wide range of step lengths are compatible with recovery from a perturbation backwards as long as V_COM_ remains low (approximately <15-20 cm/s). For V_COM_ values above 15-20 cm/s, there is a steep increase in fall risk, most prominently for trials with the smallest steps and decreasing in fall risk as step length increases.

**Figure 4.**
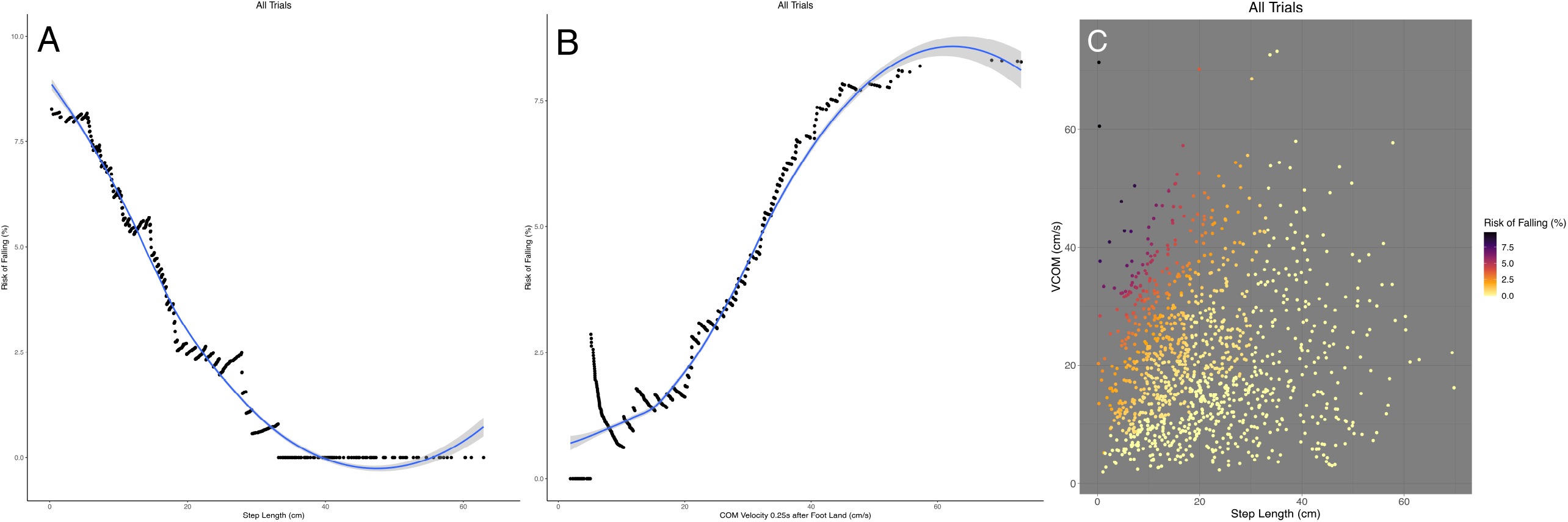
Risk of falling as a function of: A) initial step length, and B) V_COM_ 0.25s after their foot landed. C) Risk of falling illustrated as a function of the initial step length and the V_COM_ 0.25s after their foot landed. Abbreviations: V_COM_ = Center of Mass Velocity.

## Discussion

We have described a kinematic method of measuring the UPDRS_PT_ in normal pressure hydrocephalus in an effort to establish a quantitative, clinically meaningful outcome measure that can be reliably used in the clinical setting. The simplest description of the pull test’s outcome, the patient’s V_COM_ throughout the duration of the test, can reliably differentiate between UPDRS_PT_ scores within NPH patients (Figure 1A). When limited to a narrow range of perturbations to approximate a standardized pull, patients with UPDRS_PT_ scores of 2 and 3 appear quite similar (Figures 1B-C), indicating that some of the differences in V_COM_ profiles may be attributed to how hard patients are pulled. Indeed, this is typically seen as one of the disadvantages of using the UPDRS_PT_ in the clinical setting as it is difficult to standardize both between patients and providers leading to poor inter-rater reliability^6,19^. In fact, patients scored as “0” or “1” were – on average - pulled harder than those scored as “2” or “3” (Figure 1B, Supplemental Table 2), although there was significant overlap between the groups. This is likely reflective of better postural control in patients with lower scores as they were able to tolerate higher intensity pulls. Some patients demonstrated increases in mean pull intensity as their balance improved post-operatively (Supplemental Figure 3). Thus, tracking this metric over time may be a subtler indicator of clinical improvement or worsening than the UPDRS_PT_. Quantifying differences in pull intensity between providers could also serve as a possible mechanism to standardize evaluations in the future if desired.

Purposefully inducing variability into the pull test creates an advantage when investigating postural instability. Patients’ responses vary according to the intensity of the perturbation, allowing the clinician to characterize the entirety of the patient’s response (Figures 2A,2B). While clinicians often try to “standardize” their perturbation within and between patients, from a practical perspective, this task is impossible (especially when patients are seen by multiple providers).^6^ Even if the pull test (or any test of postural instability) could be standardized, our data demonstrate that a patient’s response at one standardized perturbation cannot necessarily be used as an approximation for the response at other perturbations without knowing how their response varies. Thus, depending where along the continuum of pull intensity the test was standardized, the same patient’s response could be considered adequate (not falling for a low intensity perturbation) or inadequate (falling after a high intensity perturbation). This would not change their underlying postural response, only the arbitrary manner in which we have decided to categorize them. As a result, quantifying the inherent variability present in the test is much more likely to be fruitful in terms of both understanding and classifying postural instability in movement disorder patients.

When examining their underlying postural responses by UPDRS_PT_ scores, NPH patient groups mainly differed in their overall step length and reaction time for a given intensity (y-intercept in Figures 2A,2B). Prior findings have demonstrated that the ability to scale step length with pull intensity declines with age and/or disease progression and that increased step length intercept values appear to be a compensatory measure to ensure postural stability in the face of this decline.^17^ Studies examining the postural step response using standardized perturbations in PD patients have generally shown that PD patients take smaller steps than healthy subjects,^20–23^ with slower reaction times that may be due to an increase in the number^22^ and size^21^ of anticipatory postural adjustments prior to stepping. This suggests that the kinematic deficits captured within these parameters are not necessarily disease-specific. However, one study found that the main distinction between mild and moderate PD patients was the presence of increased weight shift time and a base-width neutral step which impaired the compensatory step response but did not impact kinematic parameters.^24^ Further studies that specifically compare age- and UPDRS_PT_-matched controls to movement disorder patients with varying disease processes (e.g. PD and NPH) will need to be performed to identify age-related vs. disease-specific phenomenon.

The UPDRS_PT_ is a poor predictor of future falls in PD patients.^3,19^ This is likely due to its subjective nature, poor interrater reliability, and the coarse nature of an ordinal rating scale. In addition, the test is performed in the clinic rather than the home environment where falls typically occur, making it ecologically less valid. When analyzed in a quantitative fashion we were able to find kinematic factors (step length, pull intensity, V_COM_ 0.25s after the first step) that correlate with trials in which NPH patients “fell” in the clinic. Thus, while the UPDRS_PT_ score itself may not be a useful predictor of falling, quantitative pull test analysis may be able to identify predictors of a poor postural response that could lead to falls. This suggests some future potential applications and technique development. First, prospective trials can evaluate the utility of these kinematic parameters in predicting falls by examining a variety of patients in clinic, recording pull test kinematics and following the patients over time. Second, a “home-based” version of these parameters could be developed by sending patients home with wearable sensors in order to develop a more ecologically valid form of testing postural instability. Kinematic recording of typical home movement for patients is likely to better predict falls than clinic-based measures, provided that relevant events can be accurately captured.

This study has limitations related to its kinematic approach, technology and patient selection. Kinematics is focused on the end result of the body’s motion in space, and therefore, it gives limited insight into the underlying control mechanisms producing that result. To gain insight into the involved motor control pathways, kinematics need to be combined with other techniques such as intracranial local field potential recordings, EMG, EEG, and/or fMRI. While easily incorporated into our clinical visits, our motion capture system uses 15 IMUs and is expensive. Future work needs to be focused on reducing the relevant set of sensors to 5 or fewer that can be cheaply purchased. With regards to patient selection, this study may be limited in its generalizability as we only included patients with NPH. It is possible that patients with Parkinson’s Disease will demonstrate different kinematic patterns based on their UPDRS_PT_ score. In addition, as this study was conducted in veterans, almost all of our patients were men, meaning that our results need to be confirmed in more women.

The strengths of this approach lie in its practical advantage of incorporating the most widely used clinical test of postural instability and its analysis of the patient’s response throughout a range of perturbation intensities. Therefore, as a bridge between the clinic and laboratory, an easily adopted quantitative UPDRS_PT_ should have significantly more impact on diagnosis, monitoring, and clinical trial assessment than a test that can only be applied in certain laboratories or less widely applicable settings.

## Data Availability

Raw data were generated at the MVAHCS. Derived data supporting the findings of this study are available from the corresponding author RAM on request.

---

**Supplemental Table 1.**

| Variable | Description | Interpretation |
| --- | --- | --- |
| $V_{COM}$ | Velocity of the patients' center of mass. | Kinematic outcome. When $V_{COM} = 0$, the body has come to rest. |
| $A_{COM}$ | Acceleration of the patients' center of mass (examined only prior to step onset) | Indication of pull intensity as this is only examined prior to step onset |
| Step Length | Length of the initial step taken after the induced perturbation. | N/A |
| Slope | Slope of a linear model of Step Length v. $A_{COM}$ | Indication of how well the patient is able to scale their step to a variety of perturbation intensities. (An increased value indicates better ability to scale their step.) |
| y-Intercept | y-Intercept of a linear model of Step Length v. $A_{COM}$ | Indication of the patients' baseline step. (An increased value indicates that the participant takes a larger overall step regardless of pull intensity). |
| SLR | (Initial Step Length) / ($A_{COM}$) | Ratio of the initial step length taken to the intensity of the induced perturbation. Similar in nature to the slope of an entire pull test session, but can be calculated for each individual trial. |

Summary of variables and their descriptions. Interpretation of the variable if indicated. $V_{COM}$ = center of mass velocity. $A_{COM}$ = center of mass acceleration. SLR = step length ratio.

**Supplemental Table 2.**

| Pull Test Score Groups | Difference in Observed COM Acceleration Means (confidence interval), cm/s<sup>2</sup> | Tukey HSD p value |
| --- | --- | --- |
| 1 vs 0 | 20.9 (4.2, 37.6) | p=0.0071 |
| 2 vs 0 | -39.9 (-57, -22.8) | p<0.0001 |
| 3 vs 0 | -58.1 (-81.8, -34.4) | p<0.0001 |
| 2 vs 1 | -60.8 (-73.7, -47.9) | p<0.0001 |
| 3 vs 1 | -79 (-99.9, -58.1) | p<0.0001 |
| 3 vs 2 | -18.2 (-39.4, 3) | p=0.12 |

Observed Pull Intensities Vary between Groups.

**Supplemental Table 3.**

| Group | # of sessions | # of trials | Step length intercept |  |  |  |  | Step length slope |  |  |  |  |
| --- | --- | --- | --- | --- | --- | --- | --- | --- | --- | --- | --- | --- |
|  |  |  | Step length intercept<sup>a</sup>, p value | Group difference vs. 0 | Group difference vs. 1 | Group difference vs. 2 | Group difference vs. 3 | Step length slope<sup>a</sup>, p value | Group difference vs. 0 | Group difference vs. 1 | Group difference vs. 2 | Group difference vs. 3 |
| 0 | 8 | 133 | 23.7 (3.6), <0.0001 | -- | 9.2 (3.2), 0.02 | 18.9 (3.2), <0.0001 | 12.5 (4.1), 0.01 | 0.06 (0.02), 0.001 | -- | 0.02 (0.02), 0.72 | -0.01 (0.02), 0.94 | 0.05 (0.03), 0.14 |
| 1 | 23 | 353 | 14.5 (1.8), <0.0001 | 9.2 (3.2), 0.02 | -- | 9.7 (2.3), 0.0001 | 3.2 (3.4), 0.77 | 0.04 (0.01), <0.0001 | 0.02 (0.02), 0.72 | -- | -0.03 (0.01), 0.16 | 0.04 (0.02), 0.33 |
| 2 | 20 | 299 | 4.76 (1.2), 0.0002 | 18.9 (3.2), <0.0001 | 9.7 (2.3), 0.0001 | -- | -6.5 (3.4), 0.23 | 0.07 (0.008), <0.0001 | -0.01 (0.02), 0.94 | -0.03 (0.01), 0.16 | -- | 0.06 (0.02), 0.03 |
| 3 | 6 | 75 | 11.2 (1.6), <0.0001 | 12.5 (4.1), 0.01 | 3.2 (3.4), 0.77 | -6.5 (3.4), 0.23 | -- | 0.005 (0.01), 0.63 | 0.05 (0.03), 0.14 | 0.04 (0.02), 0.33 | 0.06 (0.02), 0.03 | -- |

Step length slope and intercept values for each group with pairwise group comparisons.
- Estimated marginal mean intercept and slope values were initially tested to see whether they were non-zero - Pairwise group comparisons were made using Tukey HSD corrections.

**Supplemental Table 4.**
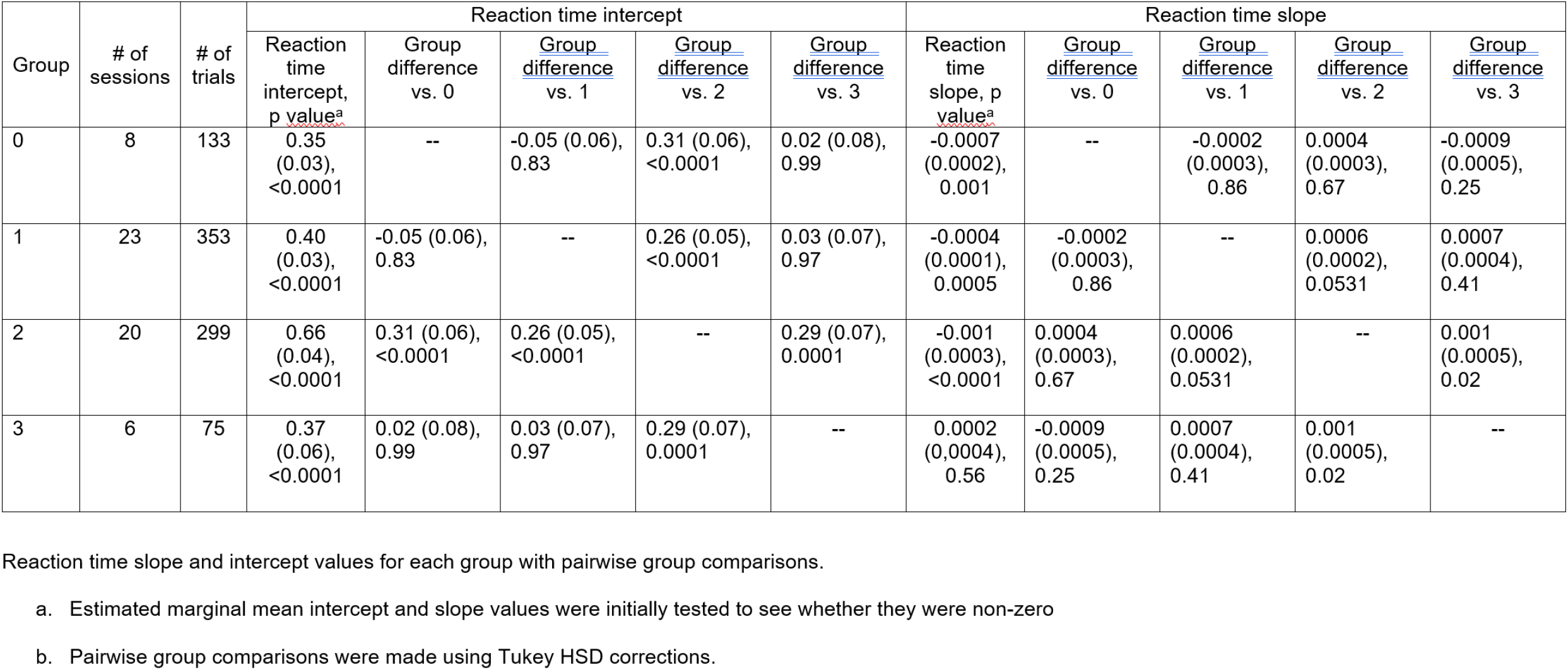

**Supplemental Figure 1.**
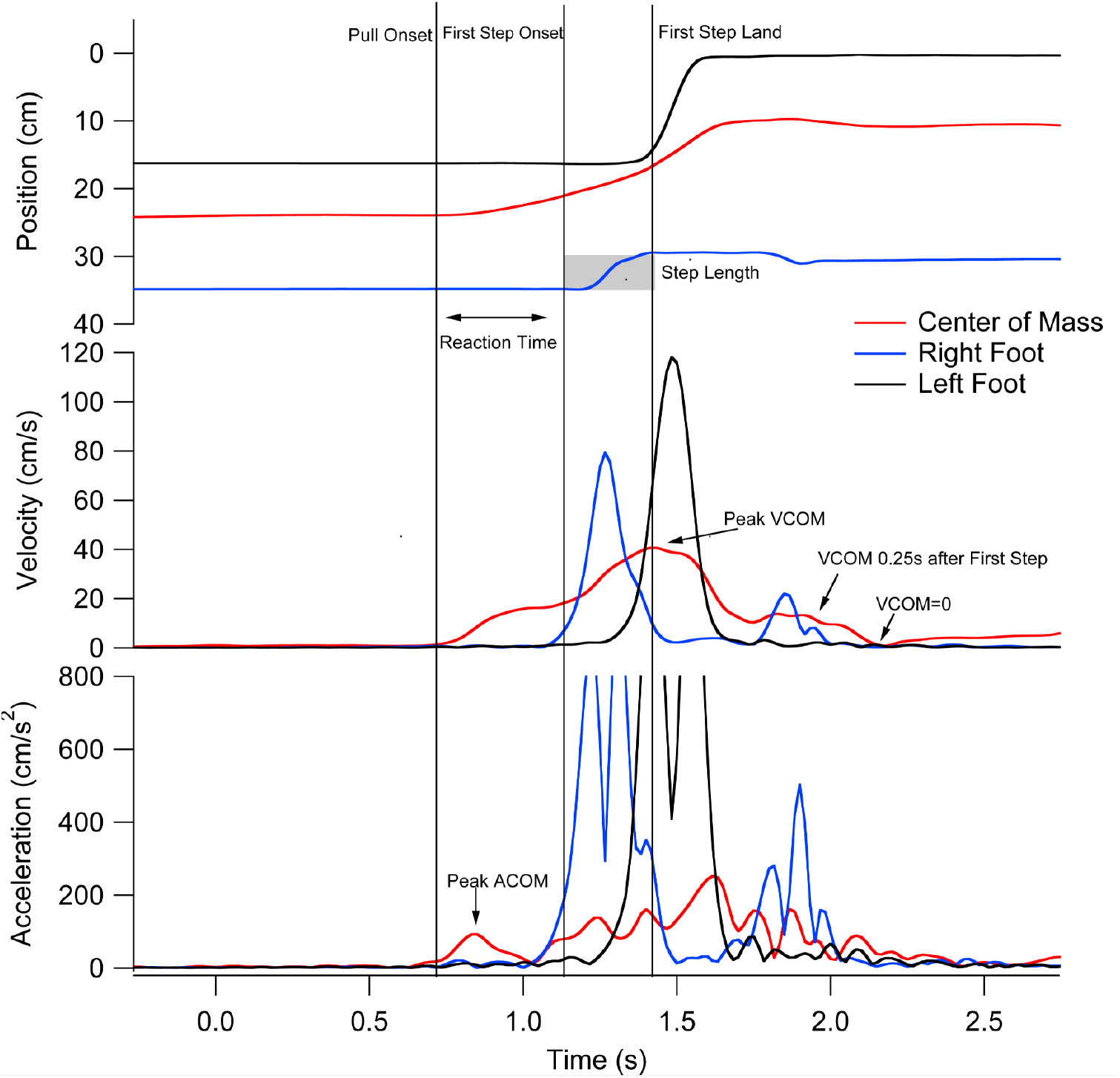
Illustration of the kinematic variables that were captured by the inertial measurement units and utilized for analysis.

**Supplemental Figure 2.**
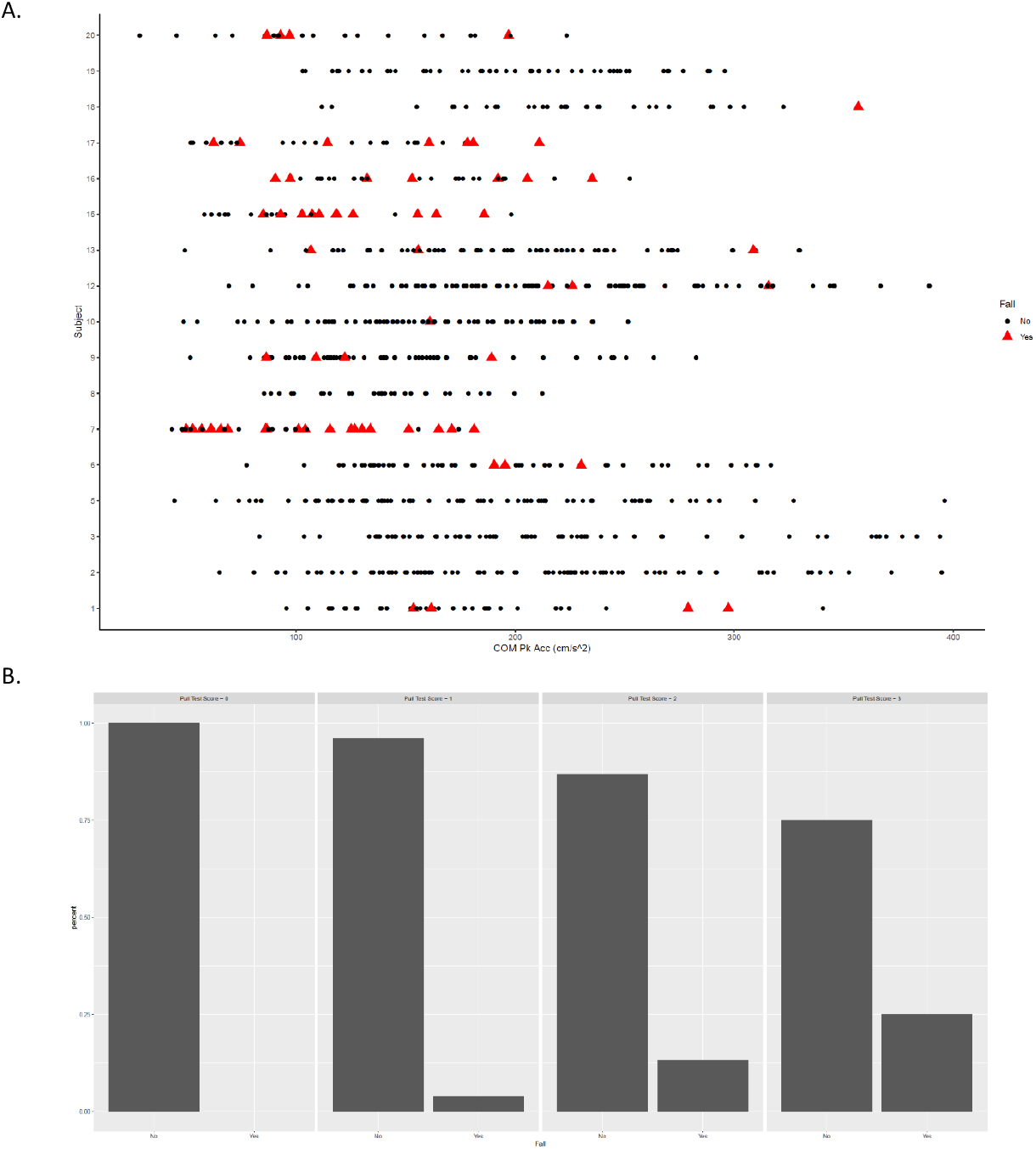
A) Illustration of the distribution of “falls” along the range of induced perturbations, stratified by patient. B) Proportion of “falls” stratified by UPDRS_PT_ score. Abbreviations: UPDRS_PT_ = Unified Parkinson’s Disease Rating Scale Pull Test. COM Pk Acc = Peak Acceleration of the Center of Mass.

**Supplemental Figure 3.**
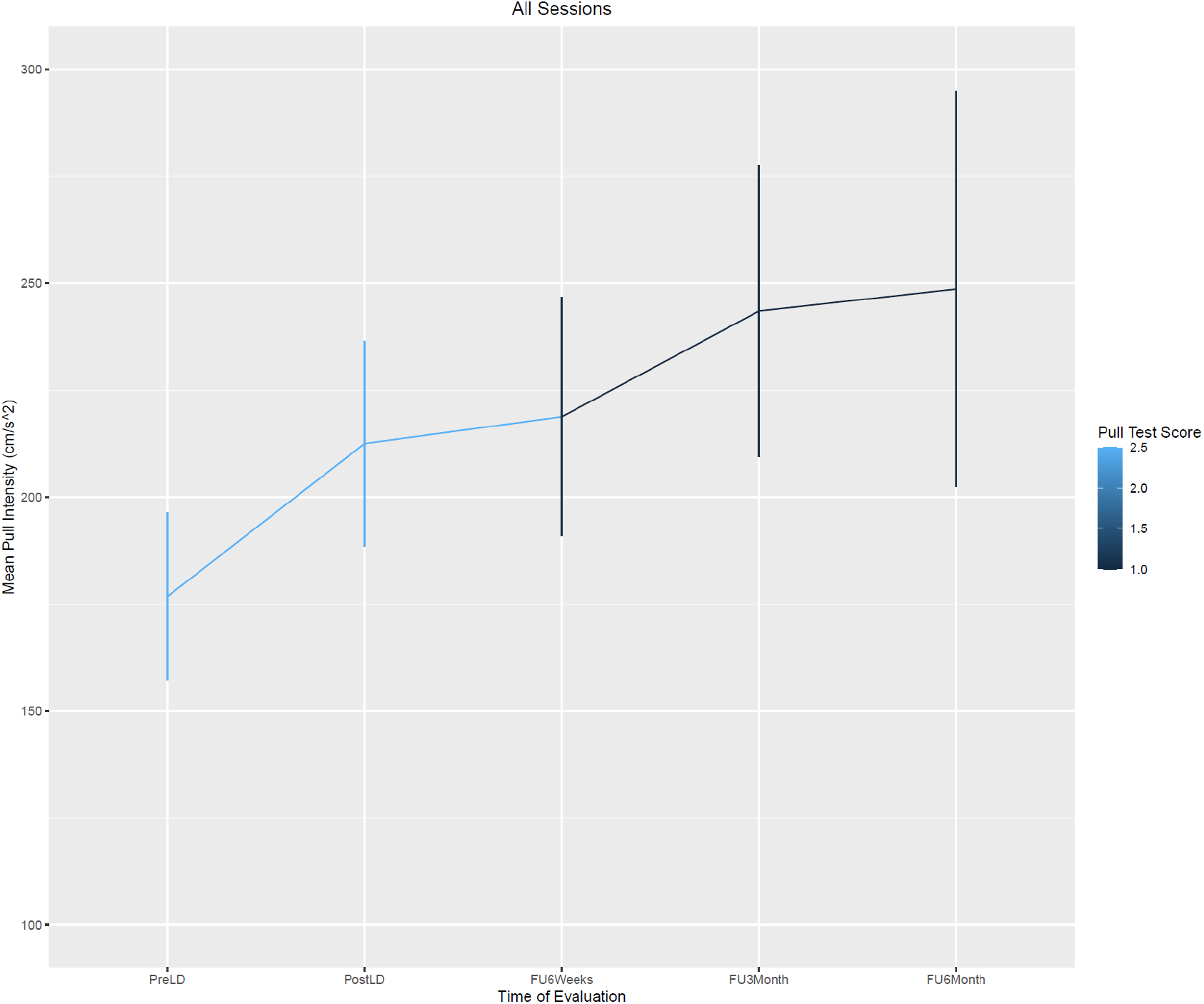
Mean pull intensity as a function of the time of evaluation. PreLD = Pre-Lumbar Drain. PostLD = Post-Lumbar Drain Treatment. FU6Weeks = 6 weeks following lumbar drain treatment. FU3Month = 3 months following lumbar drain treatment. FU6Month = 6 months following lumbar drain treatment.

